# Growth monitoring and promotion service utilization and associated factors among children under-two years of age in Samara–logia city of Afar Region, Northeast Ethiopia

**DOI:** 10.1101/2022.03.07.22272049

**Authors:** Semhal Kiros, Ibrahim Mohammed, Kedir Y. Ahmed

## Abstract

**Introduction:** Utilizing growth monitoring and promotion (GMP) during the first two years of birth helps to detect common childhood health problems (e.g., malnutrition and infections) at early stages and offers an opportunity for promoting education and nutritional counselling. However, no previously published study has investigated the level of GMP utilization and associated factors among pastoralist Ethiopian mothers including the Afar National and Regional State (ANRS), where under-nutrition is the major cause of childhood morbidity and mortality. This is the first study aimed to investigate the utilization and associated factors of GMP service for infants and young children in the ANRS of Ethiopia.

**Methods:** A community-based cross-sectional study was conducted from May to June 202 in the Samara-logia city administration. A total of 416 children aged two years were selected using a random sampling technique, and an interviewer-administered questionnaire was used to collect data. Multivariable logistic regression was applied to examine the influence of explanatory variables (including socio-demographic, obstetric and health service, and health literacy factors) on the utilization of GMP services.

**Results:** The overall utilization of GMP for infants and children was 15.9% with 95% confidence intervals (CI) from 12.0% to 19.5%. Children whose fathers attained college or higher schooling were more likely to utilize GMP services (AOR = 12.40; 95% CI: 4.15, 37.4). Children who resided in households with a higher number of children were less likely to utilize GMP services (AOR = 0.10; 95%CI: 0.03, 0.26 for households with 3-4 children and AOR = 0.16; 95% CI: 0.05, 0.48 for children with 4+). The odds for GMP service utilization was significantly higher among children who received postnatal care (AOR = 9.68; 95% CI: 3.57, 26.20).

**Conclusion:** The utilization of GMP services is lower to support the reduction of child morbidity and mortality attributed to malnutrition for infants and children in Ethiopia. A higher level of schooling and those who received postnatal care were more likely to utilize GMP, while those who were from households with a higher number of children were less likely to utilize GMP. Our findings suggest strengthening GMP service in Ethiopia, and focused efforts are required on the modifiable associated factors.

## Background

Growth monitoring and promotion (GMP) has been implemented since the 1980s worldwide with the main aims of providing a diagnostic tool to early detect health and nutritional problems of children; promoting optimal breastfeeding and appropriate complementary feeding of mothers, families and health workers; and facilitating regular contact with primary health care facilities for infants and young children[1, 2]. Theoretically, the expected benefits of GMP are achieved through improvements in the nutritional status and survival by the two chief nutritional interventions (i.e., breastfeeding and complementary feeding practices), and increases in the utilization of child health services (e.g., immunization, supplementation and referral) using a frequent health facility visits [1, 2].

Despite GMP being a standard health sector practice for global children, the impact of the program on child nutritional outcomes and survival has been a subject of debate in the past decades due to the mixed evidence of success across global nations [1-3]. For example, evidence from Nigeria, Jamaica, Madagascar, Senegal and Brazil documented the benefits of GMP [1-5], while studies from Bangladesh and India reported little or no effect of GMP [1-3, 6]. Previously published studies have documented the main barriers for the expected success of GMP: poor provider-client communication, interventions are not being tied to context-specific circumstances, skill gaps in frontline health workers (particularly in taking measurements and interpretations), shortage of time for client counselling due to patient load, and the caregivers are also not putting health advice into practice [1, 4, 6, 7]. In recognizing this fact, in 2020, the United States Agency for International Development (USAID) with other partner organizations revised the strategies of GMP to better integrate with child health services in the first 12 months of birth and to maximize the promotion element within the GMP [8].

Given the current need for maximizing the ‘promotion’ element (e.g., breastfeeding education and nutritional counselling) within the GMP and tackling the high burden of malnutrition and poor survival among children in Ethiopia [8], the Government of Ethiopia has integrated GMP with the community-based nutrition program [9, 10]. The 2016 National Nutrition Program (NNP) has also included GMP as a potential strategy for improving nutritional status and survival of under two years of age children, as part of the community-based nutrition program (CBNP) [9]. Past subnational studies in Ethiopia have also shown children who were birthed at health facilities [11], those who resided in wealthy households [11], those who lived with smaller family members [11], had mothers with good knowledge and favorable attitude [12], shorter distance to health facility [12] and those who reported antenatal use [12] were associated with higher utilization of GMP service.

Understanding what the barriers and drivers are for implementing GMP would be essential for pastoral mothers in the hottest climate Region of Afar, where the burden of malnutrition is highest due to water shortages, low agricultural production, frequent natural and manmade disasters, and poor infrastructures to access markets. Additionally, information from this study would be an input for local and global nutritional initiatives, given the 2020 NNP targets of EIBF and EBF (80% each) [9], the 2025 Ethiopian Health Sector Transformation Plan (HSTP-II) targets of stunting (reducing to 25%) [13], the 2030 Global Nutrition Targets (GNTs) of EBF (increasing to 70%) and stunting (50% reduction) [14], and the 2030 Sustainable Development Goals (SDGs) target of ending all forms of malnutrition [15]. The present study for the first time conducted to investigate the utilization and associated factors of GMP for infants and young children among pastoralist mothers in the Afar Region of Ethiopia.

## Methods

### Study design and study setting

A community-based cross-sectional study was conducted to investigate the utilization and associated factors of GMP among children under two years of age in the Samara-Logia city administration of Afar Region from May to June 2021. The ANRS is geographically located in the Great East African Rift Valley System, shares a border with Eritrea (Northeast), Tigray (Northwest), Oromia (South), Somali (Southeast), Amhara (West), and Djibouti (East) [16]. The ANRS has an estimated population size of almost 2 million with more than 12% of the population aged less than five years and the livelihood of more than 85% of the Afar population depends on livestock production (including camel, goat and cow)[17]. The arid or semi-arid and low rainfall climate makes the region vulnerable to adverse health problems including malnutrition, infection and poor survival[16].

The Samara-Logia city administration is the capital of the ANRS, located 584 km away from the capital city of Ethiopia, Addis Ababa[18], and the city has one primary hospital, two government health centers, and seven private clinics to provide preventive, curative and rehabilitative services.

### Source and study Population

The source population included all mothers/caregivers with children under two years of age who resided in the Samara-Logia city, and the study population included those who resided in the randomly selected Kebeles (the lowest administrative unit in Ethiopia context) of the administrative city.

### Sample size determination and sampling procedure

A single population proportion formula, assuming 95% confidence interval, 5% marginal error, and 43.9% proportion of GMP (based on a study that gave the optimum sample size) [19] was used to calculate the required sample. Considering a non-response rate of 10%, a total of 416 study participants were included in the study. To select the study participants, 3 out of 8 Kebeles in the Samara-Logia city were randomly (lottery method) selected, and a list of under two years children from each kebele pre-obtained from the health extension workers (community health workers in Ethiopia context) as a sampling frame. Finally, a random sampling technique that accounts for the total number of under two children in the selected Kebeles was used to select mothers/caregivers who are living with their under two years of age.

### Outcome variable

The main outcome variable for this study was GMP service utilization measured based on previously published studies in LMICS (including Ethiopia) [20, 21]. GMP utilization was defined as when a child’s height and weight measurements are recorded using a standard child growth chart at least once per month, two times per 1–3 months, five times per 4– 11 months, and four times per 12 months for 12–23 months children. For this study, the outcome variable was grouped as ‘Yes’ (GMP utilization) or ‘No’ (no GMP utilization) [20, 21].

### Explanatory variables

The explanatory variables were broadly classified as sociodemographic factors (including baby’s and mother’s age, mother’s and father’s educational status, mother’s and father’s occupation, religion, ethnicity, and wealth index), obstetric and health service factors (including ANC and PNC visits, mode of birth, place of birth, immunization and distance from a health facility), and health literacy factors (including community connectedness, awareness of GMP, and source of information). This classification was consistent with previously published studies conducted in Ethiopia and other low and middle-income countries (LMICs) [20- 23].

### Data collection and quality control

Data were collected using an interviewer-administered questionnaire designed to obtain information on explanatory factors (including sociodemographic, obstetric and health service, and health literacy factors), and the outcome variable (GMP utilization). The data collection was conducted using a Bachelor of Science Health Professionals as a data collector and a Master of Health Science Professionals as supervisor. To ensure a common understanding of the main objectives and the data collection instrument, one-day intensive training was provided by the primary investigator to the data collectors and supervisors. A pre-test was also performed on 5% of the study sample among mothers/caregivers in the Dubti District, and completeness and consistency of collected questionnaires were checked every day during the data collection period.

### Data processing and analysis

Following the collected data were checked for consistency and completeness, a data entry was conducted using an Epi-info version 7.2 software package and exported to SPSS version 27.0 for final analysis. Principal component analysis (PCA) was used to construct a household wealth index using available information on household-level assets such as television, toilet system, and source of drinking water. For this study, initial analyses involved calculating frequencies and percentages to describe the study participants and to calculate the prevalence of GMP utilization across the study participants. Multivariable logistic regression was used to examine the influence of sociodemographic, obstetric and health service, and health literacy factors on GMP. Odd ratios (ORs) and 95% confidence intervals were used to report the findings of the regression modelling, and a Hosmer and Lemeshow goodness of fit test was used for model selection.

### Ethical Consideration

Ethical clearance was obtained from the Ethical Review Committee of the College of Medical and Health Sciences of Samara University. Written informed consent was obtained from each study participant after explaining the overall objective, the confidentiality and privacy, and the expected benefits of the study and participants were reassured of the right to discontinuation at any stage during the interview process. All interviews were conducted with strict privacy in a quiet place, and the confidentiality of participants was kept by removing personal identifiers from the collected data.

## Results

### Sociodemographic factors

A total of 396 mothers/caregivers were responded in the current study, yielding a response rate of 95.0%. More than half (55.8%) of children were females, and 137 (34.6%) of them were in the age group 6-11 months. Among mothers, 181 (45.7%) of them were in the age group 25-29 years, and 290 (73.2%) of mothers were Afar in their ethnicity. More than half of mothers (53.8%) attained primary schooling or less, and nearly half (49.5%) of them resided in households with 3- 4 under-five children. The majority (81.6%) of mothers were housewives, and about 203 (51.3%) of fathers attained high school education.

**Table 1:** Sociodemographic characteristics of the respondents for GMP utilization and associated factors in the Samara-Logiya City, Afar Region, Ethiopia, 2021 (n=396)

| Variables | Frequency (n) | Percentage (%) |
| --- | --- | --- |
| **Child sex** |  |  |
| Male | 175 | 44.2 |
| Female | 221 | 55.8 |
| **Age of the child in months** |  |  |
| 0-5 months | 67 | 16.9 |
| 6-11 months | 137 | 34.6 |
| 12-17 months | 123 | 31.1 |
| 18-23 months | 69 | 17.4 |
| **Mother's age in years** |  |  |
| 15-24 years | 168 | 42.4 |
| 25-29 years | 181 | 45.7 |
| 30-34 years | 47 | 11.9 |
| **Mother's educational status** |  |  |
| Primary or less schooling | 213 | 53.8 |
| Secondary schooling | 132 | 33.3 |
| Higher education | 51 | 12.9 |
| **Mother's occupation** |  |  |
| Housewife | 323 | 81.6 |
| Government employer | 37 | 9.3 |
| Merchant | 36 | 9.1 |
| **Marital status** |  |  |
| Currently married | 369 | 93.2 |
| Formerly married | 27 | 6.8 |
| **Father's educational status** |  |  |
| Primary or less schooling | 111 | 28.0 |
| Secondary education | 203 | 51.3 |
| College/university | 82 | 20.7 |
| **Father's occupation** |  |  |
| Government employer | 179 | 45.2 |
| Private employer | 97 | 24.5 |
| Merchant | 58 | 14.6 |
| Daily laborer | 48 | 12.1 |
| Pastoralist | 14 | 3.5 |
| **Ethnicity** |  |  |
| Afar | 290 | 73.2 |
| Amhara | 60 | 15.2 |
| Tigre | 43 | 10.9 |
| Oromo | 3 | 0.8 |
| **Wealth index** |  |  |
| Poor | 173 | 43.7 |
| Middle | 98 | 24.7 |
| Rich | 124 | 31.3 |

### Health service factors

Many mothers (87.4%) reported at least one ANC visit for the current baby, while only 62 (15.7%) of them visited health facilities for PNC. The majority (85.1%) of children were birthed using normal delivery, and about 340 (85.9%) of them reported health facility birthing. One hundred eighty-three (46.2%) of mothers reported traveling more than 2- hours to reach the nearest health facility.

**Table 2:** Health service-related factors for GMP utilization and associated factors in the Samara-Logiya City, Afar Region, Ethiopia, 2021 (n=396)

| Variables | Frequency (n) | Percentage (%) |
| --- | --- | --- |
| **ANC visit** |  |  |
| Yes | 346 | 87.4 |
| No | 50 | 12.6 |
| **PNC visit** |  |  |
| Yes | 62 | 15.7 |
| No | 334 | 84.3 |
| **Mode of delivery** |  |  |
| Vaginal delivery | 337 | 85.1 |
| Operational delivery | 59 | 14.9 |
| **Place of delivery** |  |  |
| Hospital | 334 | 84.3 |
| Health center | 55 | 13.9 |
| Home | 7 | 1.8 |
| **EPI utilization** |  |  |
| Yes | 350 | 88.4 |
| No | 46 | 11.6 |
| **Distance to the nearest health facility** |  |  |
| <30 minutes | 50 | 12.6 |
| 30-60 minutes | 163 | 41.2 |
| >60 minutes | 183 | 46.2 |

### Health literacy factors

More than half of the study participants (55.8%) heard about the benefits of GMP service, and out of these, 21.7% the benefits for improving health-seeking behaviour, 15.9% reported the relevance for monitoring child growth and 7.1% reported importance for checking the health status of the child.

**Table 3:** Health literacy factors for GMP utilization and associated factors in the Samara-Logiya City, Afar Region, Ethiopia, 2021 (n=396)

| Variables | Frequency (n) | Percentage (%) |
| --- | --- | --- |
| **Heard about the benefits of GMP** |  |  |
| Yes | 175 | 44.2 |
| No | 221 | 55.8 |
| **Awareness of the benefits of GMP** |  |  |
| Seek medical care | 86 | 21.7 |
| Monitor child growth | 63 | 15.9 |
| Know health status | 28 | 7.1 |
| **Source of information on GMP** |  |  |
| Health extension workers | 30 | 7.6 |
| Other health workers | 25 | 6.3 |
| Media | 8 | 2.2 |
| **Media exposure** |  |  |
| Yes | 358 | 90.4 |
| No | 38 | 9.6 |
| **Type of media** |  |  |
| Television | 325 | 81.6 |
| Radio | 37 | 9.3 |

### Utilization of growth monitoring and promotion

The proportion of GMP utilization was 15.9% with 95% Confidence Interval [CI] from 12.0% to 19.5%, and out of this, 79.2% of them reported as regular users for the GMP services.

### Factors associated with GMP utilization

Children who resided in households with many children were less likely to utilize GMP services compared to those who resided in households with two children or less (AOR = 0.11; 95% CI: 0.05, 0.28 for households with 3-4 children and AOR = 0.24; 95% CI: 0.08, 0.67 for children with 4+ children). The odds for GMP service utilization was significantly higher among mothers who reported PNC usage for the current baby compared to those who did not report PNC use (AOR = 8.09; 95% CI: 3.19, 20.5). Children whose fathers attained college or higher education were more likely to utilize GMP services compared to those who did not have or primary schooling (AOR = 7.75; 95% CI: 3.01, 19.9).

**Table 4:**
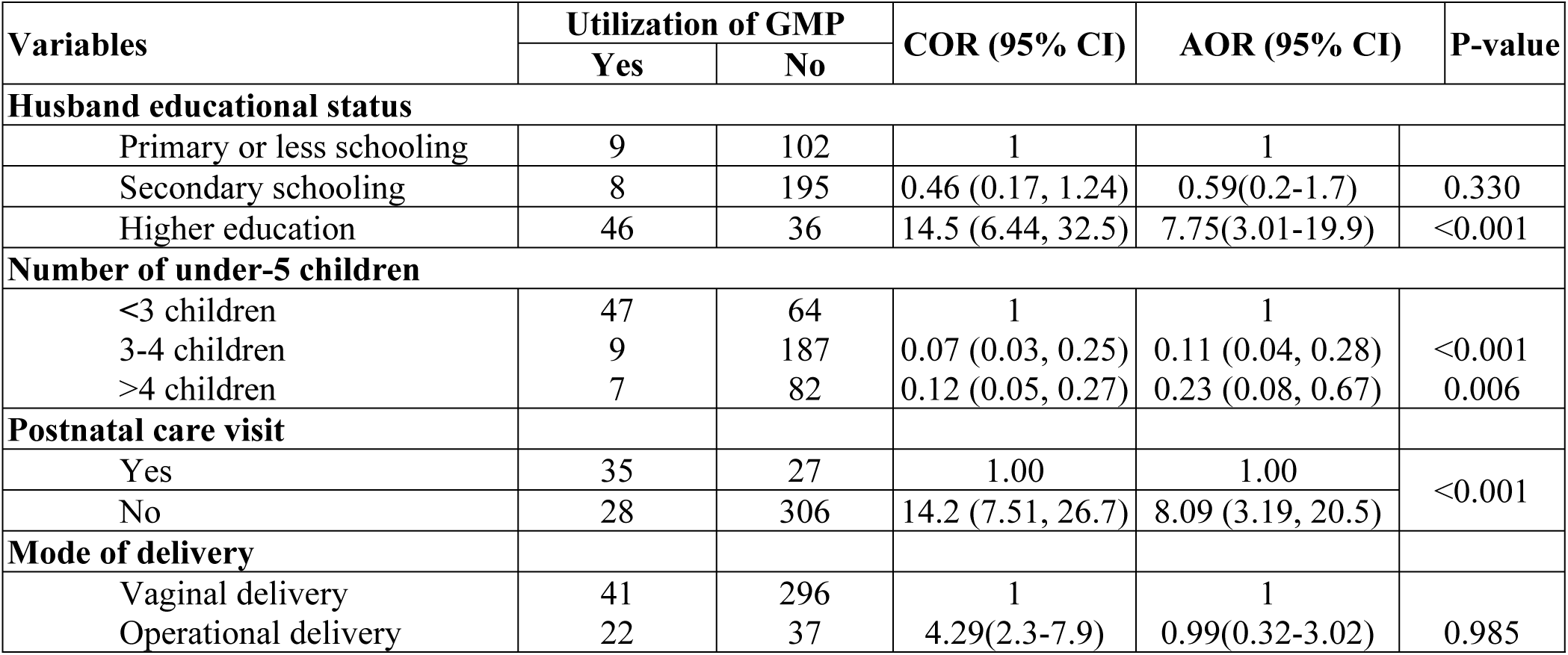
Factors associated with GMP utilization among children under two years of age in the Samara-Logiya City of Afar Region, Ethiopia, 2021 (N = 396)

## Discussion

The present study aimed to investigate the prevalence of GMP utilization and its associated factors among children under two years of age. The study showed that the overall utilization of GMP service was 15.9%, and children who resided in households with many children were less likely to utilize GMP, while those who used PNC and whose fathers attained higher education were more likely to utilize GMP.

Based on the NNP-II (2016-2020) of Ethiopia, GMP utilization was one of the strategic objectives promoted to improve the nutritional status of children under two years of age in Ethiopia. Despite this fact, the current study showed a lower level of GMP service utilization (15.9%) among under two years of age children in the Samara-Logia city of Afar Region, consistent with Ethiopian studies conducted in Mareka (16.9%) and Butajira (11.0%) districts. However, the current GMP utilization was lower as compared to the Lako Abaya study (39.0%) in Ethiopia, and other studies from South Africa (70%) [20], Rwanda (79%) [24], Kenya (53.3%) [22], Afghanistan 87% [25], Uganda (59%), Honduras (87%), Brazil (42%) and Dominican Republic (85%) [26]. The finding of the current study suggests the need for strengthening efforts for GMP utilization by maximizing the impact of the promotion element within the GMP service using evidence-based community nutrition programs.

Educated parents are less likely to seek medical care from health facilities for infants and young children, based on evidence from LMICs. The main reasons for the positive effect of parents’ education on improved child health service utilization (including GMP) are that educated parents are more like to access health-related information, with a potential improvement in the health-seeking behaviour of mothers [27, 28]. Consistent with this fact, the current study demonstrated the positive influence of the higher educational status of fathers on the utilization of GMP service for infants and young children. A qualitative study on the relationship between husband education and GMP utilization confirmed that husbands are very influential regarding decisions for health service utilization, indicating that without the support of husbands, GMP is unlikely to succeed in Ethiopia [12]. Our finding suggests the need for improving the educational status of parents to improve the impacts of GMP services in stopping the intergenerational effects of malnutrition in Ethiopia.

This study showed that children from households with a large number of under-five children were less likely to utilize GMP service in the Afar Region of Ethiopia, consistent with studies conducted in Ethiopia [21] and Indonesia [29]. The relationship between the number of children and GMP utilization can be explained in two pathways. Firstly, the increased number of children means a lower socioeconomic household. Secondly, when the number of children increases, the resources (e.g., time) of mothers/caregivers available may not allow seeking health care.

We found that mothers who visited for postnatal service were more likely to use the GMP service compared to their counterparts, consistent with a similar study conducted in the Maraka district in Ethiopia [21]. This relationship between PNC service and the higher utilization of GMP could be explained by the nutritional counselling and health education sessions provided as part of the service. The SDG target 3.8 aimed to achieve universal health coverage (UHC) to support the reduction of inequalities in health care utilization (including GMP) between well-off and disadvantaged populations [15], indicating the need to economically empower households by improving the health-seeking behaviour of mothers to detect child health problems early and provide appropriate care for infants and young children. Although the Government of Ethiopia is providing maternal and child health services free of charge in the public health facilities of Ethiopia [30], it seems that the direct and indirect costs related to time, medication and transportation are hindering the utilization of maternal and child health services (including GMP) among disadvantaged Ethiopian mothers.

The study has limitations. First, the cross-sectional nature of the study design makes it difficult to confer direction of causality, nevertheless, the findings are consistent with other studies conducted in Ethiopia [19, 21]. Second, a social desirability bias in assessing the awareness of mothers on the growth chart might be a possible limitation for the study. Despite the limitations, the study was the first to be conducted on the GMP services utilization in the pastoral community, which would support the policymakers and practitioners working in these areas.

## Conclusion

The study showed that the overall utilization of GMP service was unacceptable to improve the health, development and survival of children. Children who resided in households with many children were less likely to utilize GMP, while those who visited health facilities for PNC service and whose fathers attained higher education were more likely to utilize GMP. Our findings suggest the need for strengthening GMP service in Ethiopia, and focused efforts are required on the modifiable associated factors.

## Data Availability

All relevant data are within the manuscript and its Supporting Information files.

## Declaration

### Author’s contribution

SK conceived and designed the study, performed analysis and interpretation of data, and drafted the manuscript. KYA and IM supervised the design, conception, analysis, interpretation of data and made critical comments at each step of the research. All authors read and approved the final manuscript.

## Acknowledgments

We would like to express our sincere gratitude to the study participants, supervisors, data collectors for the commitment and time during the data collection. We are grateful to Samara University College of Medical and Health Science, Department of Public Health for setting this golden chance and providing the Master of Public Health (MPH) program.

## Ethics approval and consent to participate

The study was reviewed and approved by the ethical review committee of Samara University, College of Medical and Health Science. All participants were pre-informed about the aim of the study and their full right to withdraw or refuse to participate before their written consent was obtained

## Consent for publication

“Not applicable”

## Availability of data and materials

“The data set will not be shared to protect the participant’s identities”

